# A Machine Learning Approach to Identifying Delirium from Electronic Health Records

**DOI:** 10.1101/2021.09.09.21263247

**Authors:** Jae Hyun Kim, May Hua, Robert A. Whittington, Junghwan Lee, Cong Liu, Casey N. Ta, Edward R. Marcantonio, Terry E. Goldberg, Chunhua Weng

## Abstract

**Background:** Despite the well-known impact of delirium on long-term clinical outcomes, identification of delirium in electronic health records (EHR) remains difficult due to inadequate assessment or documentation of delirium. The purpose of this research is to present a classification model that identifies delirium using retrospective EHR data. The classification model would support the additional identification of delirium cases otherwise undocumented during routine practice.

**Methods:** Delirium was confirmed with the Confusion Assessment Method for the Intensive Care Unit (CAM-ICU). Age, sex, Elixhauser comorbidity index, drug exposures, and diagnoses were used as features to train the logistic regression and multi-layer perceptron models. The clinical notes from the EHR were parsed to supplement the features that were not recorded in the structured data. The model performance was evaluated with a 5-fold cross-validation area under the receiver operating characteristic curve (AUC).

**Results:** Seventy-six patients (17 cases and 59 controls) with at least one CAM-ICU evaluation result during ICU stay from January 30, 2018 to February 20, 2018 were included in the model. The multi-layer perceptron model achieved the best performance in identifying delirium; mean AUC of 0.967 ± 0.019. The mean positive predictive value (PPV), mean negative predicted value (NPV), mean sensitivity, and mean specificity of the MLP model were 0.9, 0.88, 0.56, and 0.95, respectively.

**Conclusion:** A simple classification model showed a mean AUC over 0.95. This model promises to identify delirium cases with EHR data, thereby enable a sustainable infrastructure to build a retrospective cohort of delirium in the ICU. The cohort would be useful for the evaluation of long-term sequelae of delirium in ICU.

## INTRODUCTION

Delirium is a frequent complication among intensive care unit (ICU) patients, with its incidence ranging between 45% and 87% of all ICU patients.[1, 2] There are short-term and long-term impacts of delirium during an ICU stay on patients’ clinical outcomes. For instance, delirium is known to be associated with prolonged hospitalization, short and long term cognitive impairment, and increased healthcare costs.[3-5] Delirium has also been associated with increased short and long-term mortality.[6-8] Nevertheless, according to ICU delirium practice guidelines, there still exists a significant research gap regarding the long-term outcomes of delirium.[3] The establishment of retrospective delirium cohorts would be useful for long-term surveillance. However, the under-coding of delirium diagnoses and the burden of delirium screening in clinical practice inhibit the identification of delirium in the electronic health records (EHR) and the establishment of retrospective cohorts.[9, 10] A number of delirium prediction models have been developed.[11, 12] Some developed multivariable models using 4-9 preoperative variables [13-15] and other recent models used machine learning or deep learning algorithms including neural net.[16, 17] However, as stated, all of them used pre-operative (or before admission) characteristics since the goal of these models was to predict ahead of time patients who may develop delirium after certain interventions such as hip surgery, in-patient admission, or ICU stay. Therefore, new diagnoses or drug prescriptions during hospitalization periods were not included in the clinical prediction model.

In contrast, the focus of this research was to retrospectively identify ICU patients who experienced delirium during hospitalization using a classification model. Considering that the occurrence of delirium would elicit a change in treatment pattern during hospitalization, the inclusion of variables recorded during hospitalization in the model could potentially increase the accuracy of the classification model. The study population included only patients who had been evaluated for delirium in the ICU using the standard Confusion Assessment Method for the Intensive Care Unit (CAM-ICU).

## METHODS

### Data

This study was approved by Columbia University Irving Medical Center (CUIMC) institutional review board and informed consent was waived. The dataset for model development included patients from either the Surgical or Cardiothoracic ICU with at least one Confusion Assessment Method for the Intensive Care Unit (CAM-ICU) evaluation result during their ICU stay at New York Presbyterian Hospital (NYP) / CUIMC from January 30, 2018 to February 20, 2018. The CAM-ICU data were obtained as a part of a quality improvement project that aimed to improve recognition of delirium in the two ICUs. Raters received training (in the form of videos and a written manual) and performed CAM-ICU assessments on a convenience sample of patients. Interrater reliability was assessed using Gwet’s kappa in a sample of 15 patients and found to be high (0.9295, 95% confidence interval (CI) 0.7689-1.000).[18] If the patient was ever positive from at least one of the CAM-ICU evaluations, that patient was counted as having post-operative delirium.

### Model Implementation

Following clinician guidance, we included the following features for model development: patients’ age at the time of admission, sex, Elixhauser comorbidity index, diagnoses (e.g., heart failure, gout, *etc*.), and drug prescription records. EHR data for these features were extracted from the Observational Medical Outcomes Partnership (OMOP) Common Data Model (CDM) version 5.3 formatted clinical data warehouse of NYP/CUIMC.[19] Different drug forms were regarded as different drug exposures, even if the drug ingredient was the same. For example, oral sodium bicarbonate and intravenous sodium bicarbonate were treated as different drug exposures. The Elixhauser comorbidity index was calculated with the records of diagnoses from six months prior to the admission to the date of admission. Age and Elixhauser comorbidity index were normalized to range from 0 to 1. Drug exposures and diagnoses were one-hot encoded (i.e., 1 denotes the presence of a drug exposure or diagnosis, while 0 denotes no drug exposure or diagnosis) and represented as a vector. A subset of EHR notes (e.g., transfer notes, delirium nurses’ notes, *etc*.) considered pertinent to ICU medical practice by clinicians was parsed. We selected notes written by the ICU team or consultants that would potentially be making assessments or recommendations for delirium. The MetaMap Lite version 3.6.2rc6 with NegEx algorithm was used in feature extraction from notes.[20, 21] The extracted concepts were normalized using the Human Phenotype Ontology[22] and further mapped to the OMOP CDM standard concepts in order to identify duplicate terms from the structured EHR data. Supplementary Table S1 lists the titles of the notes that were parsed.

We used two simple machine learning methods: logistic regression and multi-layer perceptron (MLP). We evaluated each of the models using two different inputs. The first input included the features from the structured EHR data. The second input included the phenotypes extracted from clinical notes as well as the features from the structured EHR data. The performance of each model was evaluated with 5-fold cross-validation. In each fold, the test set area under the receiver operating characteristic curve (AUC) was calculated, and the mean ± standard error of AUCs was presented. The validation set was not used in the model development. Hyperparameters were chosen using grid search based on the training loss. The number of epochs was set to 20. The learning rate was set to 0.001 for LR models and 0.0001 for MLP models. MLP models have a single hidden layer with 128 hidden units. The Adam optimizer was used in all models.[23] Python version 3.6.9 and Tensorflow version 2.3.1 were used.[24] The code is available in the following GitHub repository: https://github.com/WengLab-InformaticsResearch/delirium. The R package comorbidity was used to calculate the Elixhauser comorbidity index.[25]

## RESULTS

We identified 76 patients who were admitted to NYP/CUIMC ICU and had evaluation results for the CAM-ICU for delirium from January 30, 2018 to February 20, 2018. Table 1 shows the characteristics of patients according to the delirium status. Their mean age was 63.4 ± 15.1 years and 59.2% (n = 45) of patients were male. The mean Elixhauser comorbidity index was 4.1 ± 3.2. The mean hospitalization duration was 28 ± 36 days. The CAM-ICU evaluation was conducted 123 times in 76 patients, with 1.6 times per patient on average. Among 76 patients, 17 patients (22.4%) had delirium in at least one of the CAM-ICU evaluations. 1,318 unique features were extracted from the structured data from the 76 patients. These patients had 2,650 EHR notes of the types included in this study (Supplementary Table S1). From these notes, 657 unique concepts were extracted, 499 of which (76.0%) were only in notes. The concepts from structured data and clinical notes are listed in the Supplementary Table S2.

**Table 1.** Characteristics of patients according to the delirium evaluation results

| **Variable** | **Delirium positive**<br>**(n = 17)** | **Delirium negative**<br>**(n = 59)** | **Total**<br>**(n = 76)** |
| --- | --- | --- | --- |
| Age, mean (SD) | 68.6 (12.3) | 61.9 (15.6) | 63.4 (15.1) |
| Male, n (%) | 9 (52.9) | 36 (61.0) | 45 (59.2) |
| Elixhauser comorbidity index, mean (SD) | 5.2 (3.0) | 3.8 (3.2) | 4.1 (3.2) |
| Hospitalization duration (days), mean (SD) | 52.1 (49.9) | 21.3 (28.4) | 28.1 (36.4) |

SD, standard deviation.

Figure 1 shows the mean 5-fold cross-validation AUC of all evaluated models. The MLP model showed the highest mean AUC (0.967 ± 0.019), followed by MLP+notes (0.965 ± 0.032), LR+notes (0.962 ± 0.038), and LR (0.957 ± 0.014). Models with note concepts had higher standard deviation than models without notes. Figure 2 shows the receiver operating characteristic curve (ROC) of all evaluated models. When the threshold of the MLP model was 0.81, mean positive predictive value (PPV), mean negative predictive value (NPV), mean sensitivity, and mean specificity were 0.9, 0.88, 0.56, and 0.95, respectively. Figure 3 shows the precision-recall curve of all evaluated models. The precision-recall curve of the MLP model was well above the baseline of 0.22 (proportion of positive cases among evaluated samples).

**Figure 1.**
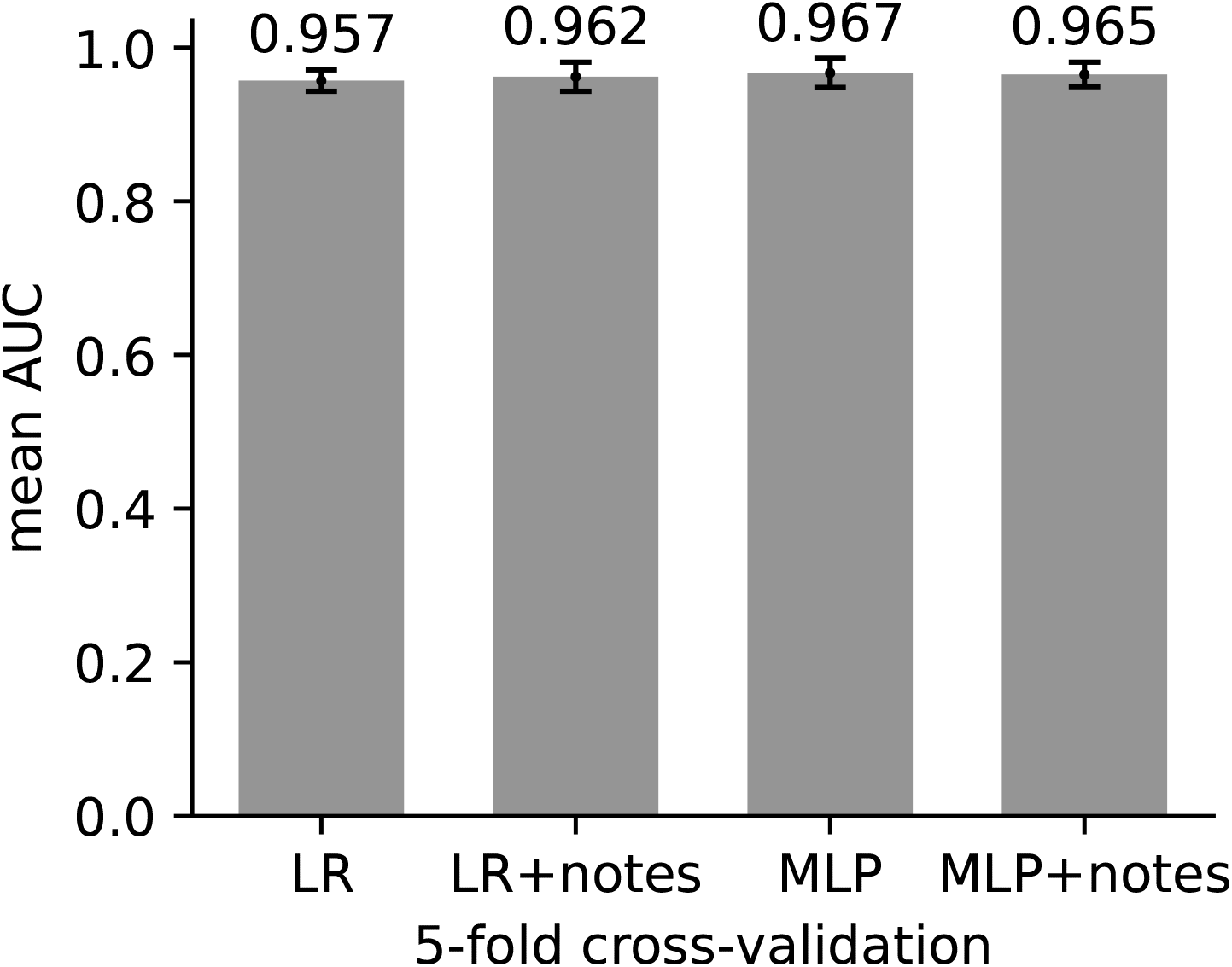
Mean 5-fold cross validation AUC of all models. AUC, area under the receiver operating characteristic curve; LR, logistic regression; MLP, multi-layer perceptron

**Figure 2.**
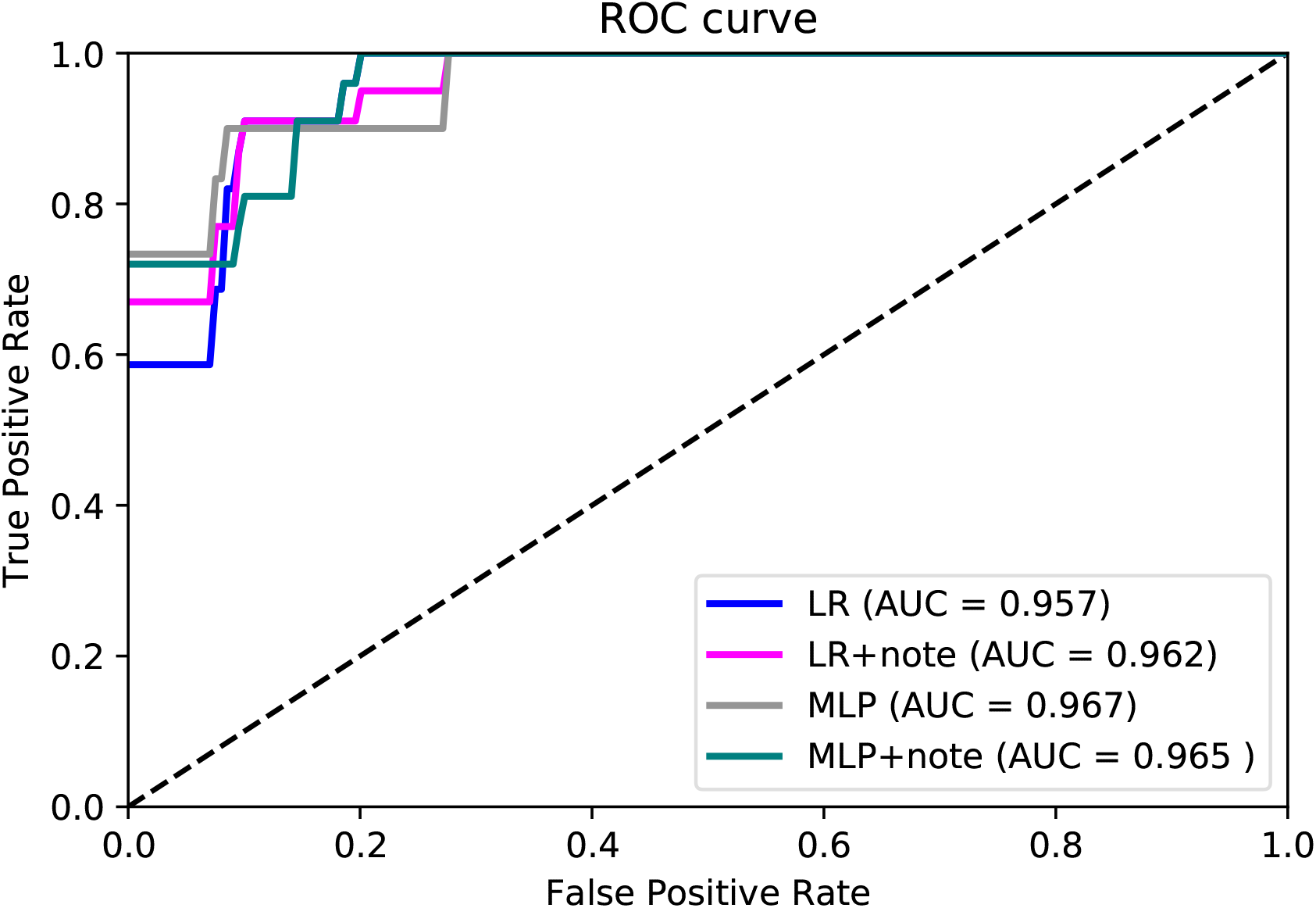
ROC curve of all models. ROC, receiver operating curve; LR, logistic regression; MLP, multi-layer perceptron

**Figure 3.**
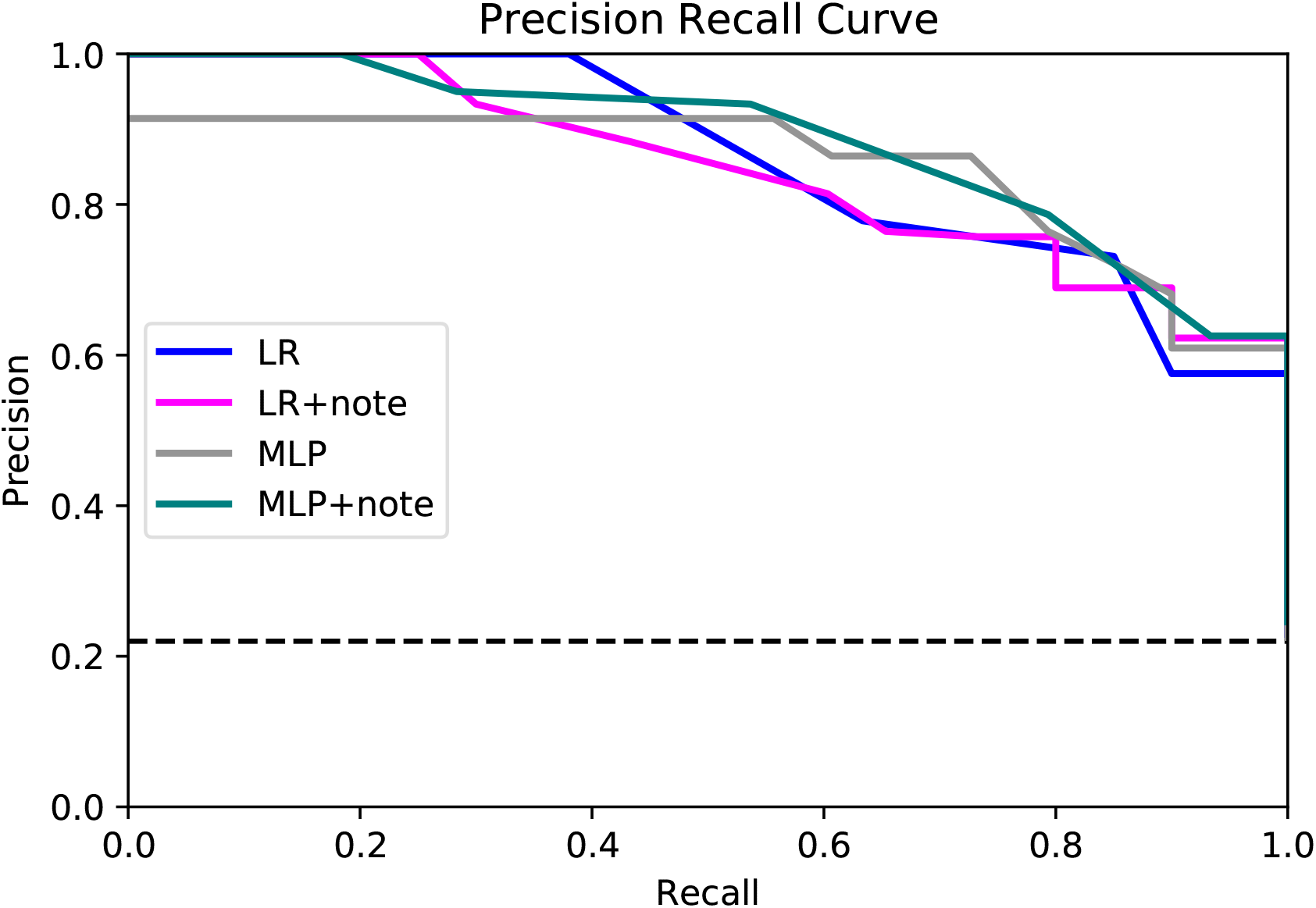
Precision-recall curve of all models. LR, logistic regression; MLP, multi-layer perceptron. Dash line shows the proportion of positive cases among evaluated sample (0.22). All curves were well above the baseline.

## DISCUSSION

We applied the logistic regression and MLP model to classify ICU patients into delirium cases or controls using retrospective EHR data. We expect our models to have potential in the identification of patients with delirium in ICU while minimizing the human effort to manually review the accumulated patients’ records. Such a model would be useful for patient follow-up for better determining the long-term medical sequelae of ICU patients with a history of delirium.

A few clinical prediction models have been developed to predict delirium either prospectively or retrospectively.[11, 12] Previous studies focused more on the development of predictive models rather than classification models. Therefore, existing predictive models used features mostly at baseline and did not fully incorporate the diagnosis or drug prescription during admission even with retrospective data. One study that used features during admission also limited the drug prescription records that were extracted 1 day before the diagnosis of delirium.[26] The higher AUC of our model can be attributed to the inclusion of patients’ comorbidities before and during admission thereby comprehensively incorporating clinical status into the model. Besides the higher mean AUC of the model, another advantage is that our model used the CAM-ICU to evaluate the patients as delirium positive or negative, while others used chart-based methods.[26, 27] The CAM-ICU is the recommended screening tool for delirium in ICU according to the practice guideline.[3]

We also compared the model with features extracted from the EHR notes with the model without the notes-derived concepts. Interestingly, inclusion of these additional concepts did not lead to improvement of model performance. Possible reasons include low frequency of each concept documented in notes. If more patients were included in the training data set, additional data from the unstructured EHR notes could add more value to the model. In addition, automated natural language processing using MetaMap Lite is known to have lower precision and recall when compared to manual review of clinical notes.[28] The use of automated natural language processing may have introduced noise into the model.

Our classification model would be useful in the identification of missed patients with delirium and thereby augments the clinical diagnosis of delirium in combination with patient screening at bedside, albeit retrospectively. Patients with delirium could be under-documented due to multiple reasons. First, factors including misinterpretation of patients’ status, documentation errors, and education gap may lead to inappropriate documentation of CAM-ICU evaluation results.[29, 30] Second, in clinical practice, not all patients in the ICU receive the CAM-ICU evaluation for delirium, despite quality improvement efforts.[31] Third, according to the study by Chanques *et al*., CAM-ICU had a sensitivity of 83%, and among the 108 CAM-ICU ratings, seven cases were false negatives.[32] For these reasons, a subset of delirium patients remains unevaluated, undocumented, or otherwise unidentified and hence lose a chance at being followed for the occurrence of complications of the delirium. Moreover, the evaluation of delirium was even more difficult in the recent COVID-19 pandemic.[33] The COVID-19 survivors who had ICU care may possess high-risk for long-term cognitive sequelae, with a recent multi-center study finding a delirium prevalence of 54.9%.[34]

This study has limitations. First, our model only included a small number of patients (n = 76) compared to previous studies that developed machine learning algorithms to classify delirium. Because of the small sample size, we used 5-fold cross validation to evaluate model performance instead of using a held-out test set to prevent overfitting. The small amount of data can also restrict developing or applying complex machine learning models that have a lot of parameters to train. Second, some information that is relevant to a comprehensive delirium evaluation, including neurological examination results (verbal, motor, and eye response), performance statuses, or magnetic resonance imaging (MRI) reports of the brain (FLAIR [Fluid-Attenuated Inversion Recovery] signal intensity, hippocampal atrophy, *etc*.), were not systematically available from the whole patient cohort. Third, the biomarkers that have recently been studied as having an association with delirium were not included as features in our model. Our cohort consisted of patients in early 2018 and the levels of biomarkers including tau, interleukin 8, and neurofilament light (NfL) protein were not available in most of the patients, as these tests are not yet routinely performed in a clinical setting. Further research is required to evaluate the role of these biomarkers in the classification model. Also, this study is subject to the traditional limitations of observational data. Finally, the portability of this model to other EHR data in other institutions should be further tested.

## Conclusion

We present a classification model that identifies patients with a delirium episode during their ICU stay using retrospective data. The classification model showed high accuracy with a mean AUC over 0.95. The model could be used in the retrospective identification of undiagnosed delirium cases and the establishment of a delirium cohort for long-term evaluation and surveillance.

## Data Availability

The datasets generated and/or analyzed during the current study are not publicly available due to patient privacy.

## Acknowledgements

The authors would like to thank Kenrick D. Cato and Sarah C. Rossetti for reviewing this paper.

## Author contributions

JHK wrote the manuscript; JHK, MH, TEG, RAW, and CW designed the research; MH provided the CAM-ICU evaluation results; MH selected the notes in the electronic health records for parsing; JL prepared the machine learning codes; JHK, JL, CL, and CNT refined the machine learning algorithm; JHK analyzed the data; CL contributed to the natural language processing of notes in the electronic health records; JHK, MH, RAW, JL, CL, CNT, ERM, TEG, and CW edited and approved the manuscript.

## Ethics approval and consent to participate

This study was approved by Columbia University Irving Medical Center (CUIMC) institutional review board and informed consent was waived.

## Consent for publication

Not applicable.

## References

1. Ely EW, Margolin R, Francis J, May L, Truman B, Dittus R, Speroff T, Gautam S, Bernard GR, Inouye SK: Evaluation of delirium in critically ill patients: validation of the Confusion Assessment Method for the Intensive Care Unit (CAM-ICU). Crit Care Med 2001, 29(7):1370–1379.

2. Roberts B, Rickard CM, Rajbhandari D, Turner G, Clarke J, Hill D, Tauschke C, Chaboyer W, Parsons R: Multicentre study of delirium in ICU patients using a simple screening tool. Aust Crit Care 2005, 18(1):6, 8-9, 11–14 passim.

3. Devlin JW, Skrobik Y, Gelinas C, Needham DM, Slooter AJC, Pandharipande PP, Watson PL, Weinhouse GL, Nunnally ME, Rochwerg B et al: Clinical Practice Guidelines for the Prevention and Management of Pain, Agitation/Sedation, Delirium, Immobility, and Sleep Disruption in Adult Patients in the ICU. Crit Care Med 2018, 46(9):e825–e873.

4. Goldberg TE, Chen C, Wang Y, Jung E, Swanson A, Ing C, Garcia PS, Whittington RA, Moitra V: Association of Delirium With Long-term Cognitive Decline: A Meta-analysis. JAMA Neurol 2020.

5. Kinchin I, Mitchell E, Agar M, Trepel D: The economic cost of delirium: A systematic review and quality assessment. Alzheimers Dement 2021.

6. Witlox J, Eurelings LS, de Jonghe JF, Kalisvaart KJ, Eikelenboom P, van Gool WA: Delirium in elderly patients and the risk of postdischarge mortality, institutionalization, and dementia: a meta-analysis. JAMA 2010, 304(4):443–451.

7. Chalmers LA, Searle SD, Whitby J, Tsui A, Davis D: Do specific delirium aetiologies have different associations with death? A longitudinal cohort of hospitalised patients. Eur Geriatr Med 2021.

8. Bai J, Liang Y, Zhang P, Liang X, He J, Wang J, Wang Y: Association between postoperative delirium and mortality in elderly patients undergoing hip fractures surgery: a meta-analysis. Osteoporos Int 2020, 31(2):317–326.

9. Eastwood GM, Peck L, Bellomo R, Baldwin I, Reade MC: A questionnaire survey of critical care nurses’ attitudes to delirium assessment before and after introduction of the CAM-ICU. Aust Crit Care 2012, 25(3):162–169.

10. Hope C, Estrada N, Weir C, Teng CC, Damal K, Sauer BC: Documentation of delirium in the VA electronic health record. BMC Res Notes 2014, 7:208.

11. Lindroth H, Bratzke L, Purvis S, Brown R, Coburn M, Mrkobrada M, Chan MTV, Davis DHJ, Pandharipande P, Carlsson CM et al: Systematic review of prediction models for delirium in the older adult inpatient. BMJ Open 2018, 8(4):e019223.

12. Ruppert MM, Lipori J, Patel S, Ingersent E, Cupka J, Ozrazgat-Baslanti T, Loftus T, Rashidi P, Bihorac A: ICU Delirium-Prediction Models: A Systematic Review. Crit Care Explor 2020, 2(12):e0296.

13. Kim EM, Li G, Kim M: Development of a Risk Score to Predict Postoperative Delirium in Patients With Hip Fracture. Anesth Analg 2020, 130(1):79–86.

14. Rudolph JL, Jones RN, Levkoff SE, Rockett C, Inouye SK, Sellke FW, Khuri SF, Lipsitz LA, Ramlawi B, Levitsky S et al: Derivation and validation of a preoperative prediction rule for delirium after cardiac surgery. Circulation 2009, 119(2):229–236.

15. Marcantonio ER, Goldman L, Mangione CM, Ludwig LE, Muraca B, Haslauer CM, Donaldson MC, Whittemore AD, Sugarbaker DJ, Poss R et al: A clinical prediction rule for delirium after elective noncardiac surgery. JAMA 1994, 271(2):134–139.

16. Davoudi A, Ebadi A, Rashidi P, Ozrazgat-Baslanti T, Bihorac A, Bursian AC: Delirium Prediction using Machine Learning Models on Preoperative Electronic Health Records Data. Proc IEEE Int Symp Bioinformatics Bioeng 2017, 2017:568–573.

17. Mufti HN, Hirsch GM, Abidi SR, Abidi SSR: Exploiting Machine Learning Algorithms and Methods for the Prediction of Agitated Delirium After Cardiac Surgery: Models Development and Validation Study. JMIR Med Inform 2019, 7(4):e14993.

18. De Raadt A, Warrens MJ, Bosker RJ, Kiers HAL: Kappa Coefficients for Missing Data. Educ Psychol Meas 2019, 79(3):558–576.

19. Voss EA, Makadia R, Matcho A, Ma Q, Knoll C, Schuemie M, DeFalco FJ, Londhe A, Zhu V, Ryan PB: Feasibility and utility of applications of the common data model to multiple, disparate observational health databases. J Am Med Inform Assoc 2015, 22(3):553–564.

20. Chapman WW, Bridewell W, Hanbury P, Cooper GF, Buchanan BG: A simple algorithm for identifying negated findings and diseases in discharge summaries. J Biomed Inform 2001, 34(5):301–310.

21. Demner-Fushman D, Rogers WJ, Aronson AR: MetaMap Lite: an evaluation of a new Java implementation of MetaMap. J Am Med Inform Assoc 2017, 24(4):841–844.

22. Kohler S, Gargano M, Matentzoglu N, Carmody LC, Lewis-Smith D, Vasilevsky NA, Danis D, Balagura G, Baynam G, Brower AM et al: The Human Phenotype Ontology in 2021. Nucleic Acids Res 2021, 49(D1):D1207-D1217.

23. Kingma DP, Ba J: Adam: A Method for Stochastic Optimization. CoRR 2014, abs/1412.6980.

24. Abadi M, Barham P, Chen JM, Chen ZF, Davis A, Dean J, Devin M, Ghemawat S, Irving G, Isard M et al: TensorFlow: A system for large-scale machine learning. In: Proceedings of Osdi’16: 12th Usenix Symposium on Operating Systems Design and Implementation. Berkeley: Usenix Assoc; 2016: 265–283.

25. Gasparini A: comorbidity: An R package for computing comorbidity scores. Journal of Open Source Software 2018, 3(23):648.

26. de Wit HA, Winkens B, Mestres Gonzalvo C, Hurkens KP, Mulder WJ, Janknegt R, Verhey FR, van der Kuy PH, Schols JM: The development of an automated ward independent delirium risk prediction model. Int J Clin Pharm 2016, 38(4):915–923.

27. Rudolph JL, Doherty K, Kelly B, Driver JA, Archambault E: Validation of a Delirium Risk Assessment Using Electronic Medical Record Information. J Am Med Dir Assoc 2016, 17(3):244–248.

28. Liu C, Peres Kury FS, Li Z, Ta C, Wang K, Weng C: Doc2Hpo: a web application for efficient and accurate HPO concept curation. Nucleic Acids Res 2019, 47(W1):W566-W570.

29. Riekerk B, Pen EJ, Hofhuis JG, Rommes JH, Schultz MJ, Spronk PE: Limitations and practicalities of CAM-ICU implementation, a delirium scoring system, in a Dutch intensive care unit. Intensive Crit Care Nurs 2009, 25(5):242–249.

30. Terry KJ, Anger KE, Szumita PM: Prospective evaluation of inappropriate unable-to-assess CAM-ICU documentations of critically ill adult patients. J Intensive Care 2015, 3:52.

31. Stewart C, Bench S: Evaluating the implementation of confusion assessment method-intensive care unit using a quality improvement approach. Nurs Crit Care 2018, 23(4):172–178.

32. Chanques G, Ely EW, Garnier O, Perrigault F, Eloi A, Carr J, Rowan CM, Prades A, de Jong A, Moritz-Gasser S et al: The 2014 updated version of the Confusion Assessment Method for the Intensive Care Unit compared to the 5th version of the Diagnostic and Statistical Manual of Mental Disorders and other current methods used by intensivists. Ann Intensive Care 2018, 8(1):33.

33. Rebora P, Rozzini R, Bianchetti A, Blangiardo P, Marchegiani A, Piazzoli A, Mazzeo F, Cesaroni G, Chizzoli A, Guerini F et al: Delirium in Patients with SARS-CoV-2 Infection: A Multicenter Study. J Am Geriatr Soc 2021, 69(2):293–299.

34. Pun BT, Badenes R, Heras La Calle G, Orun OM, Chen W, Raman R, Simpson BK, Wilson-Linville S, Hinojal Olmedillo B, Vallejo de la Cueva A et al: Prevalence and risk factors for delirium in critically ill patients with COVID-19 (COVID-D): a multicentre cohort study. Lancet Respir Med 2021, 9(3):239–250.

