## supplementary table s1 for "A Machine Learning Approach to Identifying Delirium from Electronic Health Records"

**Supplementary Table S1.** The list of parsed electronic health records notes.

| **Note title** | **Number of notes** | **Number of patients with notes** |
| --- | --- | --- |
| CTICU/SICU Daily Progress Note | 1,627 | 76 |
| CTICU Free Text Note | 199 | 56 |
| Transfer Note | 130 | 66 |
| Palliative Care Follow-Up Note | 88 | 11 |
| CCU Progress Note | 85 | 9 |
| Medical ICU Progress Note | 84 | 2 |
| Medical Critical Care Attending ICU Note | 81 | 2 |
| Psychiatry Consult Follow Up - Resident/Fellow Note | 66 | 8 |
| Medicine Resident Progress Note | 66 | 7 |
| Surgical Intensive Care Unit (SICU) Resident Note | 54 | 27 |
| Psych Consultant Note | 52 | 13 |
| Delirium Nurses Note | 28 | 15 |
| Neurology Consultation Note | 22 | 18 |
| Medical Student Follow-Up Free Text Note | 16 | 5 |
| Palliative Care Initial Consultation | 14 | 13 |
| Psychiatry Consult Initial - Resident/Fellow Note | 8 | 8 |
| Palliative Care Follow-Up Note-Free Text | 8 | 6 |
| Medical Critical Care Attending ICU Admission Note | 6 | 3 |
| Medical ICU Admission Note | 4 | 3 |
| Medical Student Admission Free Text Note | 3 | 2 |
| Psychiatry Consult Follow Up - Medical Student Note | 2 | 1 |
| Surgical Intensive Care Unit (SICU) Attending Note | 2 | 2 |
| Psychiatry Consult Initial - Medical Student Note | 1 | 1 |
| Medical Critical Care ICU/Triage Attending Consult Note | 1 | 1 |
| Medical Critical Care Stepdown Unit Consult Note | 1 | 1 |
| Medicine Admission Free Text Note | 1 | 1 |
| Medicine Admission Semi-Structured Note | 1 | 1 |

CTICU, cardiothoracic intensive care unit; SICU, surgical intensive care unit; CCU, cardiac care unit; ICU, intensive care unit
